# Real-time PCR assay for detection and differentiation of *Coccidioides immitis* and *Coccidioides posadasii* from culture and clinical specimens

**DOI:** 10.1101/2021.04.08.21254780

**Authors:** Sudha Chaturvedi, Tanya R. Victor, Anuradha Marathe, Ketevan Sidamonidze, Kelly L. Crucillo, Vishnu Chaturvedi

**Author notes:** Species-specific real time PCR for *C. immitis* and *C. posadasii. Lugar Center for Public Health Research, 0184 Tbilisi, Georgia (Ketevan Sidamonidze).

## Abstract

Coccidioidomycosis (Valley Fever) is a pulmonary and systemic fungal disease with increasing incidence and expanding endemic areas. The differentiation of etiologic agents *Coccidioides immitis* and *C. posadasii* remains problematic in the clinical laboratories as conventional PCR and satellite typing schemes are not facile. Therefore, we developed Cy5- and FAM-labeled TaqMan-probes for duplex real-time PCR assay for rapid differentiation of *C. immitis* and *C. posadasii* from culture and clinical specimens. The *RRA2* gene encoding proline-rich antigen 2, specific for *Coccidioides* genus, was the source for the first set of primers and probe. *Coccidioides immitis* contig 2.2 (GenBank: AAEC02000002.1) was used to design the second set of primers and probe. The second primers/probe did not amplify the corresponding *C. posadasii* DNA, because of an 86-bp deletion in the contig. The assay was highly sensitive with limit of detection of 0.1 pg gDNA/PCR reaction, which was equivalent to approximately ten genome copies of *C. immitis* or *C. posadasii*. The assay was highly specific with no cross-reactivity to the wide range of fungal and bacterial pathogens. Retrospective analysis of fungal isolates and primary specimens submitted from 1995 to 2020 confirmed 129 isolates and three primary specimens as *C. posadasii* and 23 isolates as *C. immitis* from human coccidioidomycosis cases, while all eight primary samples from two animals were confirmed as *C. posadasii*. A preliminary analysis of cerebrospinal fluid (CSF) and pleural fluid samples showed positive correlation between serology tests and real-time PCR for two of the 15 samples. The *Coccidioides* spp. duplex real-time PCR will allow rapid differentiation of *C. immitis* and *C. posadasii* from clinical specimens and further augment the surveillance of coccidioidomycosis.

## INTRODUCTION

Coccidioidomycosis (Valley Fever) is a fungal disease caused by two closely related pathogens: *Coccidioides immitis* and *C. posadasii. Coccidioides immitis* is endemic to the San Joaquin Valley of California. *Coccidioides posadasii* is found in the desert regions of the southwestern United States, including Arizona, Utah, New Mexico, and West Texas, and parts of Mexico, Argentina, Paraguay, and Central America (1). *Coccidioides immitis* is likely to present outside the recognized endemic area as evident from human infections and DNA-positive soil samples from Washington state (2, 3).

The prevalence of *Coccidioides* infection has increased over the years; in year 2011, more than 20,000 cases were reported in the US, twice as many cases as tuberculosis (4). Further documented increase in the number of cases in California and Arizona were published in the recent updates (5, 6). It is believed that the fungus infects more than 150,000 people per year and many of whom are sick without knowing the cause or have cases so mild that they are not detected (4). The rise in *Coccidioides* infection is attributed to increased travel, relocation to endemic areas, and possible broader distribution of *C. immitis* and *C. posadasii* than previously recognized (7). Arthroconidia produced by these fungi are highly infectious, and climate change, including dry and hot weather followed by dust storms makes these conidia easily air borne to cause pulmonary infection (8). The lung infections typically resolves rapidly leaving the patient with a robust acquired immunity to re-infection (9). However, in some individuals, the disease may progress to a chronic pulmonary condition or to a systemic disease involving the meninges, bones, joints, subcutaneous, and cutaneous tissues (10). Identifying Coccidioides to the species level is likely beneficial in the proper treatment of the patients and disease surveillance.

*Coccidioides immitis* and *C. posadasii* are not easily differentiated in the clinical laboratory on account of similar morphology. *Coccidioides posadasii* was first differentiated from *C. immitis* by microsatellite analysis (11, 12). However, this approach was time consuming and technically challenging. Subsequently, several real-time PCR assays, based on LightCycler and TaqMan chemistries, were developed to differentiate *C. immitis* from *C. posadasii* (13, 14). Both approaches rely on a single nucleotide polymorphism in different regions of the target gene, and it may have limitations when large number of strains of *C. immitis* and *C. posadasii* are tested. Umeyama et al (15) described disparity PCR using species-specific primers designed from the Ci45815 PCR fragment (GenBank AB597180). Contiguous deletion of 86-bp nucleotides in corresponding *C. posadasii* contig Cp 45910 PCR fragment (GenBank AB597183) resulted in convenient distinction of *C. immitis* from *C. posadasii* by conventional PCR. We describe a TaqMan duplex real time PCR assay using *PRA2* gene encoding proline-rich antigen specific for the *Coccidioides* genus and *C. immitis* PCR fragment Ci45815 specific for *C. immitis*. The first set of primers and probe targets the *PRA2* gene specific for *Coccidioides* genus. The second set of primers and probe targets *C. immitis* contig (CiC) 2.2 (GenBank: AAEC02000002.1). The duplex real-time PCR assay was compared against redesigned diagnostic conventional PCR with 100% concordance for culture isolates. We show that duplex real-time PCR assay is highly sensitive (10 gene copies) and identifies *C. immitis* and *C. posadasii* from culture and primary specimens. The reliable, rapid and sensitive duplex real-time PCR assay will help diagnose and track coccidioidomycosis, supplementing travel history, and obviating the need for complex genotyping assays.

## MATERIALS AND METHODS

### *Coccidioides* isolates and primary specimens

One hundred fifty two isolates of *Coccidioides* species, 10 formalin-fixed paraffin-embedded tissues, three vials of Rhesus monkey kidney (RhMK) cell (Lot # A491216B) (16), two cerebrospinal fluid (CSF), two bronchial wash, and one whole blood sample, received as a part of reference service from 1995- to 2020, from various diagnostic laboratories of the New York State and neighboring states’ laboratories were part of this investigation. Thirteen known isolates of *C. immitis* and four known isolates of *C. posadasii* were tested when one of us (VC) implemented this test at the Microbial Diseases Laboratory, California State Department of Public Health. All cultures were stored as water and glycerol stocks at 30° C and −80° C, respectively. Fifteen CSF and pleural fluids, which tested positive for coccidioidomycosis by immunodiffusion and complement fixation at the University of California Davis, were also included in the study.

### DNA extraction

Extraction of DNA from *Coccidioides* spp. was carried out in the biological safety cabinet-2 (BSC-2) in the biological safety level 3 (BSL 3) laboratory, and DNA extraction from primary human and animal specimens was carried out in BSC-2 in the BSL 2 laboratory. Qiagen DNA minikit on the Qiacube automated extractor was used for all DNA extraction. In brief, approximately 5 x 5 mm size of *Coccidioides* fungal mat grown on SDA slant for 7 to 10 days was removed by sterile loop and suspended in lysis buffer containing approximately 0.2 g of glass beads and incubated for one h at 90°C. The heat-killed fungal mat was transferred to the BSL2 laboratory and homogenized in the Precellys homogenizer at 6,500 rpm for 15•sec each time (program number 5; 6500-3×60-015). The homogenized suspension was transferred to a 2-ml screw-cap tube, leaving behind the beads. DNA from homogenized samples was extracted using the Qiagen DNA mini kit in the QiaCube semiautomated DNA extractor, resulting in 50•µl of eluted DNA. For the RhMK cell line, approximately 2 ml medium containing fungal growth was first centrifuged at 12,000 RPM; the supernatant was decanted, and fungal pellet was processed for DNA extraction as described above. DNA from paraffin-embedded tissue was prepared by first sectioning the samples, and then the paraffin was dissolved with 1•ml of xylene, followed by two washes using 1•ml of 100% ethanol. The sample was then dried and extracted using the Qiagen DNA mini kit as described above with an incubation temperature of 70°C instead of 90°C. For other primary samples (blood, CSF, bronchial wash, and pleural fluid), approximately 400 μl of each samples was added to DNA extraction buffer containing beads, heated at 56 ^°^ C for 10 min followed by bead beating and DNA extraction in QiaCube as described above. All extracted DNAs were stored at −80°C. DNA of fungal species (yeasts and molds) other than *Coccidioides* spp. were procured from the Wadsworth Center Mycology Laboratory (WCML) DNA Collection Repository (Supplementary Table 1). DNA from bacterial species included *Bacillus megaterium, Escherichia coli, Nocardia farcinica, Pseudomonas aeruginosa*, and *Streptococcus pneumonia* was procured from the Wadsworth Center Bacteriology Laboratory.

### Modified conventional PCR

A modification of conventional diagnostic PCR described by Umeyama et al (15) was used. The primers were designed from the conserved region of Ci45815 (GenBank No. AB597180.1) and Cp45810 (GenBank No. AB597183.1) flanking the deleted region of *C. posadasii*. The nucleotide sequences of the diagnostic primers were V2119 5’-CCGGGTACTCCGTACATCAC-3’, and V2120 5’-ATGCGTGAAGCCAATTCTTT-3’ and the PCR conditions were initial denaturation at 95°C for 1 min followed by 30 cycles consisted of denaturation at 94°C for 1 min, annealing at 55°C for 1 min, and extension at 68°C for 1 min followed by final extension at 68°C for 3 min.

### Duplex Real-Time PCR Assay

The *Prp2* gene and contig Ci41815 (15) were used to design the duplex real-time PCR assay. The nucleotide sequences of primers and probe for *PRA2* gene target were forward primer V1753 5’GTGCGAGAAGTTGACCGACTT-3’, reverse primer V1754 5’AGGCGTGATCTTTCCTGGAA-3’, probe V1755 5’-Cy5’-AAGTGCCACTGCGCCAAGCCC-3BHQ and primers and probe for contig Ci41815 target were forward primer V2116 5’-GGTGAAATGCCCGAAAAGAG-3’ reverse primers V2118 5’-CCAATCCTTAGGTAACCGTGAG-3’, and probe V2117 5’/56FAMTTGCACTTT/ZEN/CGTTGACTAGCCGC/3IABkFQ/-3’. Reaction for each real-time PCR contained 1× PerfeCTa multiplex qPCR ToughMix (Quanta Biosciences), a 1,000•nM concentration of primers and a 250•nM concentration of probes, and 2••l of genomic DNA (approximately 1 to 10•ng) from isolates or 5 μl of tissue DNA extracted from primary clinical specimens in a final volume of 20 μl. Each PCR run also included 2••l (1•ng) of positive extraction control (C735; *C. posadasii*), 2••l (1•ng) of positive amplification controls (C735, *C. posadasii* and 249, *C. immitis*), and 2••l of negative extraction (extraction reagents only) and negative amplification (sterilized nuclease-free water) controls. Parallel to each PCR assay, inhibitory PCR was also performed by incorporating 1 ng of *Coccidioides* gDNA into each primary clinical DNA sample. The unidirectional workflow kept the reagent preparation, specimen preparation, and amplification and detection areas separate to avoid cross-contamination. Cycling conditions on the ABI 7500 FAST system (Applied Biosystems, Thermo Fisher Scientific Inc., Waltham, MA) were initial denaturation at 95°C for 20 s, followed by 45 cycles of 95°C for 3 s and 60°C for 30 s. Based on the limit of detection (LOD), a cycle threshold (*C*_*T*_) value of •38 was reported as positive; and >38 was reported as negative. For all primary samples, specimens were reported as inconclusive if PCR inhibition was observed for the primary specimens.

### Analytical sensitivity, specificity, and reproducibility of the duplex real-time PCR assay

The 10-fold serial dilutions of genomic DNA from one isolate each of *C. posadasii* (C-735) and *C. immits* (249) were used to assess the analytical sensitivity of the duplex real-time PCR assay. The assay specificity was assessed by using an extensive DNA panel comprising various fungi both closely and distantly related to *Coccidioides* spp. and few bacterial pathogens primarily responsible for causing pulmonary infection (Supplementary Table 1). The assay reproducibility was determined by using varying concentration of gDNA from three isolates each of *C. posadasii* and *C. immitis*, ran on three different days in duplicate (inter-assay reproducibility), and on the same day in triplicate (intra-assay reproducibility). The assay’s precision was assessed using blinded panel with varying concentration of gDNA from 20 isolates each of *C. posadassii* and *C. immitis*, and 10 isolates of fungi other than *Coccidioides* spp.

### Statistical analysis

The results were statistically analyzed using GraphPad Prism 8.0 software for macOS. The statistical significance was set at a *P* value of < 0.05. All the Ct values were averages of at least three repetitions for sensitivity, reproducibility assays and average of two or three Ct values for duplex real-time PCR assay.

## RESULTS

### Modified conventional PCR assay

Initially, we modified conventional diagnostic PCR assay using species-specific primers designed from the Ci45815 PCR fragment (GenBank AB597180) described by Umeyama et al (15). The modification reduced the amplicon length down to 200-bp for *C. immitis* and 114-bp for *C. posadasii*, yielding better electrophoretic separation (Figure 1). Also, PCR efficiency was higher, with a smaller amplicon size (data not shown). Of 152 isolates investigated, 129 were *C. posadasii*, and 23 were *C. immitis* by conventional PCR. This approach was not successful for the analysis of *Coccidioides* DNA from primary specimens except for RhMK cell samples (Table 1). Overall, our results showed that the conventional diagnostic PCR could be used successfully to identify *C. immitis* and *C. posadasii* from culture isolates but not from the primary specimens.

**Table 1.** Detection of *Coccidioides* species DNA from primary human and animal specimens

| Sample type | Source | Culture | Histopathology | Diagnostic PCR (bp) | Duplex Real-Time PCR |  | Final ID |
| --- | --- | --- | --- | --- | --- | --- | --- |
|  |  |  |  |  | Mean Ct (Cy5) | Mean Ct (FAM) |  |
| Whole blood | Human (case 1) | Negative | NA | Negative | 0 | 0 |  |
| Tongue tissue (PB) | Human (case 2) | NA | Positive | Negative | 0 | 0 | Negative |
| Tongue tissue (PB) | Human (case 2) | NA | Positive | Negative | 31.86 | 0 | *C. posadasii* |
| Lung tissue (PB) | Human (case 3) | NA | NA | Negative | 34.45 | 0 | *C. posadasii* |
| Tissue (PB) | Human (case 4) | NA | NA | Negative | 37.58 | 0 | *C. posadasii* |
| Tissue (PB) | Human (case5) | NA | NA | Negative | 31.84 | 0 | *C. posadasii* |
| Cerebrospinal fluid | Human (case 6) | Negative | NA | Negative | 0 | 0 | Negative |
| Cerebrospinal fluid | Human (case 7) | Negative | NA | Negative | 0 | 0 | Negative |
| Bronchial wash | Human (case 8) | Negative | NA | Negative | 0 | 0 | Negative |
| Trachobronchial lymph node (PB) | Rhino (case 1) | NA | Positive | Negative | 32.44 | 0 | *C. posadasii* |
| Left lung (PB) | Rhino (case 1) | NA | Positive | Negative | 35.72 | 0 | *C. posadasii* |
| Left liliac lymph node (PB) | Rhino (case 1) | NA | Positive | Negative | 36.86 | 0 | *C. posadasii* |
| Left Supramammary lymph node (PB) | Rhino (case 1) | NA | Positive | Negative | 34.82 | 0 | *C. posadasii* |
| Synoviam (PB) | Rhino (case 1) | NA | Positive | Negative | 34.42 | 0 | *C. posadasii* |
| RhMK cell vial 1 | Monkey (case 1) | Positive | NA | 114 | 18.56 | 0 | *C. posadasii* |
| RhMK cell vial 2 | Monkey (case 1) | Positive | NA | 114 | 19.35 | 0 | *C. posadasii* |
| RhMK cell vial 3 | Monkey (case 1) | Positive | NA | 114 | 22.15 | 0 | *C. posadasii* |

NA = Not available; PB = Paraffin block

**Fig. 1.**
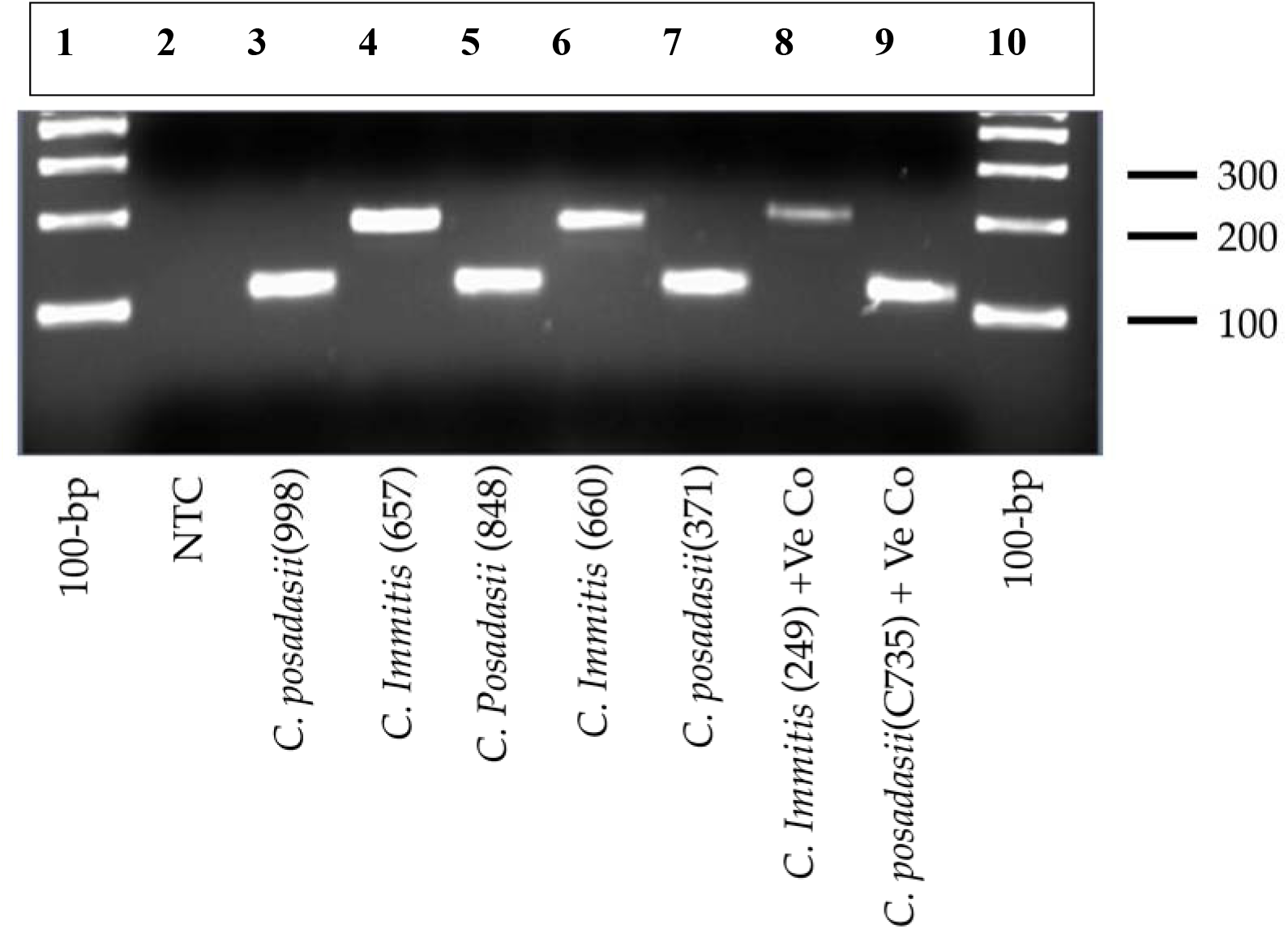
A modified conventional diagnostic PCR assay. A modification of assay described by Umeyama et al. (2006) allowed generation of smaller amplicons while maintaining test accuracy. Lane 1, and 10; 100-bp DNA ladder; lane 2 non-template control (NTC), lanes 3, 5, 7, and 9 *C. posadasii*; lanes 4, 6, 8 *C. immitis*. The identity of the isolates was also confirmed by Sanger sequencing.

### Duplex real-time PCR assay sensitivity, specificity, and reproducibility

We designed a duplex real-time PCR assay with the first set of primers and probe targeting *PRA2* gene identifying *Coccidioides* species, and the second set of primers and probe targeting *C. immitis* contig Ci41815 (CiC) identifying *C. immitis* but not *C. posadasii* due to 86-bp deletion in the corresponding *C. posadasii* contig Cp41810. The duplex real-time PCR assay was highly sensitive, with the limit of detection was 0.1 pg gDNA/PCR reaction, which was equivalent to approximately ten genome copies of *C. immitis* or *C. posadasii* (Figure 2, and Table 2). None of the other fungal or bacterial DNA yielded any Ct value, confirming the high specificity of the duplex real-time PCR assay (Supplementary Table 1). The assay was highly reproducible as it yielded correct ID for *C. immitis* and *C. posadasii* when varying concentration of gDNA tested within the same day or on different days (Supplementary Table 2 A and 2 B), and also when samples were blinded by one operator and tested by another operator (Supplementary Table 3). Since *Coccidioides* genus comprises only two species, positive results with both probes identified *C. immitis* while the positive, negative results with *PRA2* and Ci41815 probes, respectively, identified *C. posadasii* (Table 3).

**Table 2.** Duplex real-time PCR Assay Sensitivity

| gDNA dilutions<br>ng/PCR reaction | Probe | *C. posadasii* |  |  | *C. immitis* |  |  |
| --- | --- | --- | --- | --- | --- | --- | --- |
|  |  | Ct 1 | Ct 2 | Mean Ct | Ct 1 | Ct 2 | Mean Ct |
| 100 | *PRA2 (Cy5)* | 18.66 | 18.86 | 18.76 | 19.04 | 19.14 | 19.09 |
| 10 |  | 22.36 | 22.28 | 22.32 | 23.16 | 23.58 | 23.16 |
| 1 |  | 26.34 | 25.96 | 26.15 | 27.00 | 27.17 | 26.88 |
| 0.1 |  | 29.80 | 30.05 | 29.92 | 30.83 | 30.75 | 30.63 |
| 0.01 |  | 33.21 | 33.47 | 33.34 | 33.97 | 33.69 | 34.12 |
| 0.001 |  | 36.38 | 35.25 | 35.81 | 37.37 | 36.65 | 37.01 |
| 0.0001 |  | 0.0 | 0.0 | 0.0 | 0.0 | 0.0 | 0.0 |
| 100 | *CiC (FAM)* | 0.0 | 0.0 | 0.0 | 19.40 | 19.47 | 19.43 |
| 10 |  | 0.0 | 0.0 | 0.0 | 23.58 | 23.50 | 23.54 |
| 1 |  | 0.0 | 0.0 | 0.0 | 27.17 | 27.40 | 27.28 |
| 0.1 |  | 0.0 | 0.0 | 0.0 | 30.75 | 31.40 | 31.07 |
| 0.01 |  | 0.0 | 0.0 | 0.0 | 33.69 | 34.78 | 34.23 |
| 0.001 |  | 0.0 | 0.0 | 0.0 | 37.83 | 37.50 | 37.66 |
| 0.0001 |  | 0.0 | 0.0 | 0.0 | 0.0 | 0.0 | 0.0 |

**Table 3.** Test interpretation of real-time PCR assay for *Coccidioides* species

| Test Interpretation | Probe |  |
| --- | --- | --- |
|  | *Prp2* (Cy5) | *CiC* (FAM) |
| *Coccidioides immitis* | Positive | Positive |
| *Coccidioides posadasii* | Positive | Negative |
| Other Fungi | Negative | Negative |
| Bacteria | Negative | Negative |

**Fig. 2.**
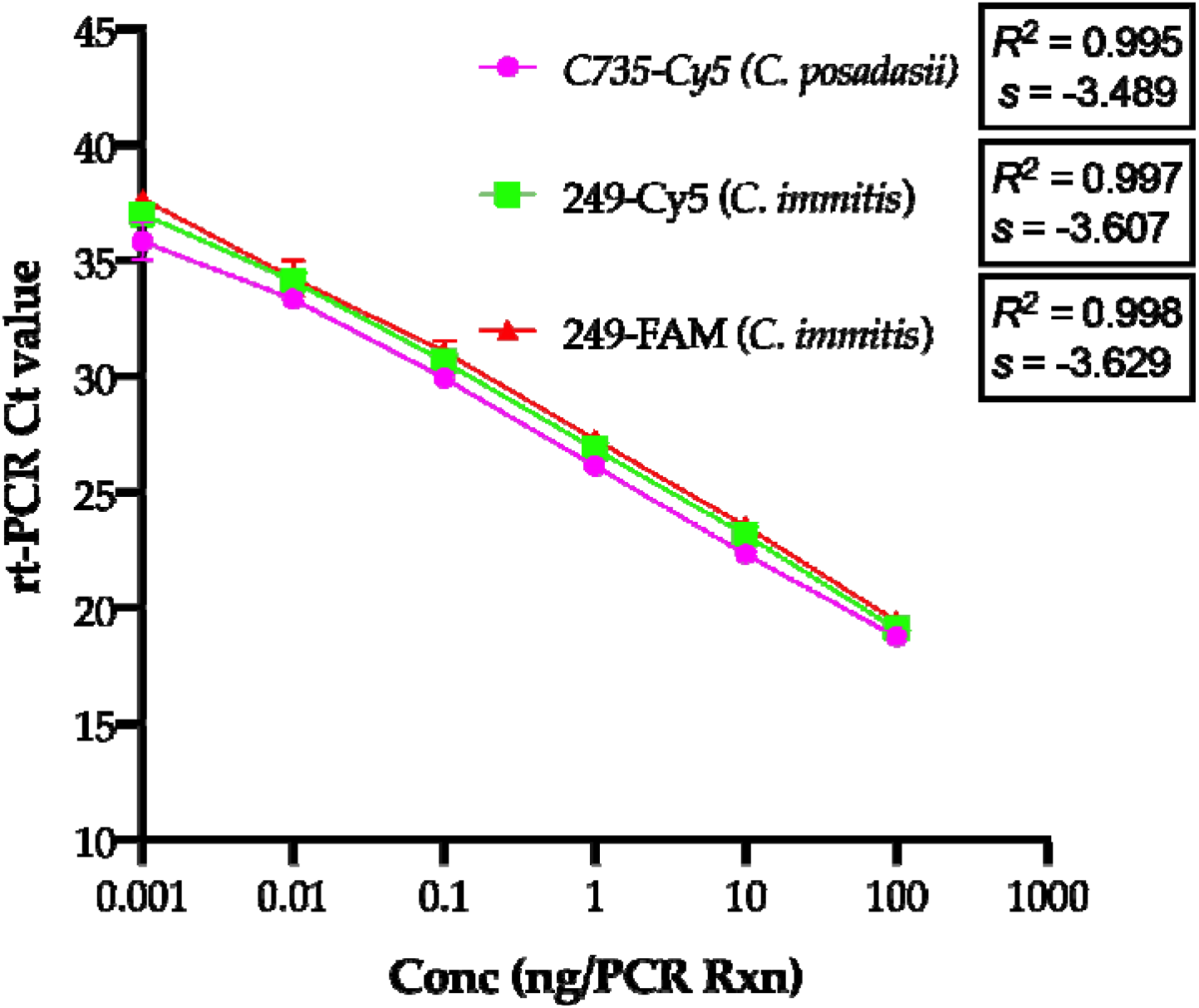
Coccidioides spp. duplex real-time PCR assay sensitivity. Genomic DNA from two control strains of *Coccidioides species, C. immitis* (249) and *C. posadasii* (C-735) were serially diluted and tested in duplicate in duplex real-time PCR assay. The assay was linear over 6 orders of magnitude and the limit of detection was 0.001 ng or 1 picogram gDNA /PCR reactions at 45 PCR cycles confirming high sensitivity. As indicated, the assay targeting *CiC* gene did not yield Ct values against *C. posadasii* DNA. The test was repeated with similar results.

### Performance of the duplex real-time PCR assay

The duplex real-time PCR assay correctly identified all cultures of *Coccidioides* species to either *C. immitis* or *C. posadasii*, and the results were corroborated with modified diagnostic PCR assay, confirming the high utility of the duplex real-time PCR assay. Of 152 human isolates of *Coccidioides* received for reference testing, 129 were identified as *C. posadasii*, and 23 were identified as *C. immitis* (Fig. 3). Of 17 isolates received from California, 13 were confirmed as *C. immtits* and four were confirmed as *C. posadasii* (Table 4). Next, we assessed the duplex real-time PCR assay’s ability to detect *Coccidioides* species DNA from 17 primary specimens. Nine of ten paraffin-embedded tissues and three vials of RhMK cells were positive for *C. posadassi* DNA, while all other samples were negative for *Coccidioides* DNA. The clinical samples negative for *Coccidioides* DNA were also negative for any other fungal DNA by internal transcribed spacer (ITS) PCR and for fungal pathogens by culture (Table 1). All of the three RhMK cell vials positive for C. *posadasii* DNA also yielded *Coccidioides* in culture, and subsequently confirmed as *C. posadasii* (Table 1). We also evaluated 15 archived CSF and pleural fluid samples, which were earlier tested positive for coccidoidomycosis by immunodiffusion and complement fixations tests. Of these samples, only two were positive for *C. posadasii* DNA by duplex real-time PCR assay (Table 5).

**Table 4.** Results of real-time PCR testing of *Coccidioides* spp. isolates from California

| No. | Initial identification | Conventional PCR (size in bp) | Duplex Real-Time PCR |  | Final Interpretation |
| --- | --- | --- | --- | --- | --- |
|  |  |  | Mean Ct (Cy5) | Mean Ct (FAM) |  |
| 497-97 | *C. immitis* | 200 | 24.33 | 31.09 | *C. immitis* |
| 498-97 | *C. immitis* | 114 | 20.19 | 0 | *C. posadassii*<sup>#</sup> |
| 662-98 | *C. immitis* | 200 | 23.02 | 22.25 | *C. immitis* |
| 664-98 | *C. immitis* | 200 | 32.24 | 32.61 | *C. immitis* |
| 665-98 | *C. immitis* | 114 | 24.74 | 0 | *C. posadassii*<sup>#</sup> |
| 666-98 | *C. immitis* | 114 | 18.76 | 0 | *C. posadassii*<sup>#</sup> |
| 667-98 | *C. immitis* | 200 | 29.8 | 28.66 | *C. immitis* |
| 669-98 | *C. immitis* | 114 | 27 | 0 | *C. posadassii*<sup>#</sup> |
| 243-09 | *C. immitis* | 200 | 18.56 | 19.98 | *C. immitis* |
| 244-09 | *C. immitis* | 200 | 28.59 | 30.18 | *C. immitis* |
| 245-09 | *C. immitis* | 200 | 26.2 | 27.6 | *C. immitis* |
| 246-09 | *C. immitis* | 200 | 21.81 | 20.87 | *C. immitis* |
| 247-09 | *C. immitis* | 200 | 25.17 | 25.81 | *C. immitis* |
| 248-09 | *C. immitis* | 200 | 36.08 | 37.73 | *C. immitis* |
| 249-09 | *C. immitis* | 200 | 17.62 | 17.73 | *C. immitis* |
| 250-09 | *C. immitis* | 200 | 19.92 | 21.28 | *C. immitis* |
| 251-09 | *C. immitis* | 200 | 22.95 | 21.74 | *C. immitis* |

<sup>#</sup> Confirmed by Sanger sequencing.

**Table 5.**
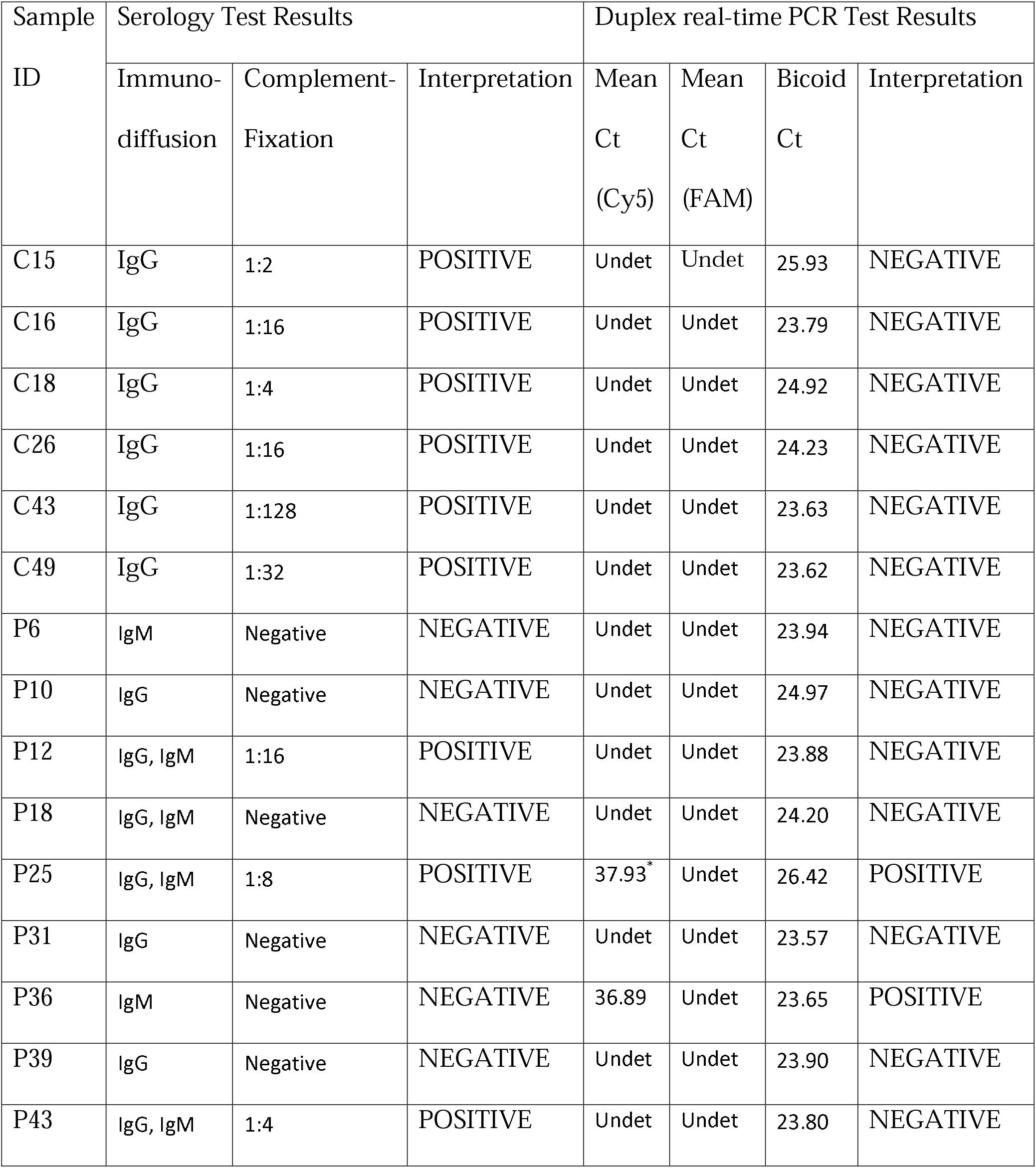
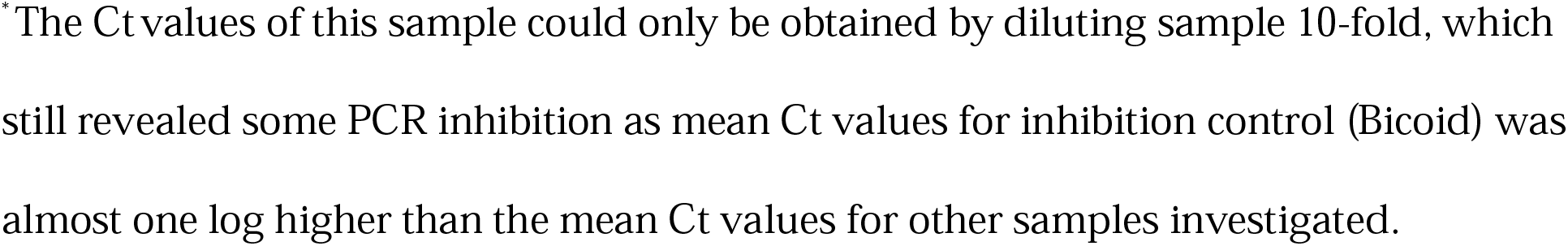
Evaluation of archived CSF and pleural fluid samples by real time PCR for *Coccidioides species*.

**Fig. 3.**
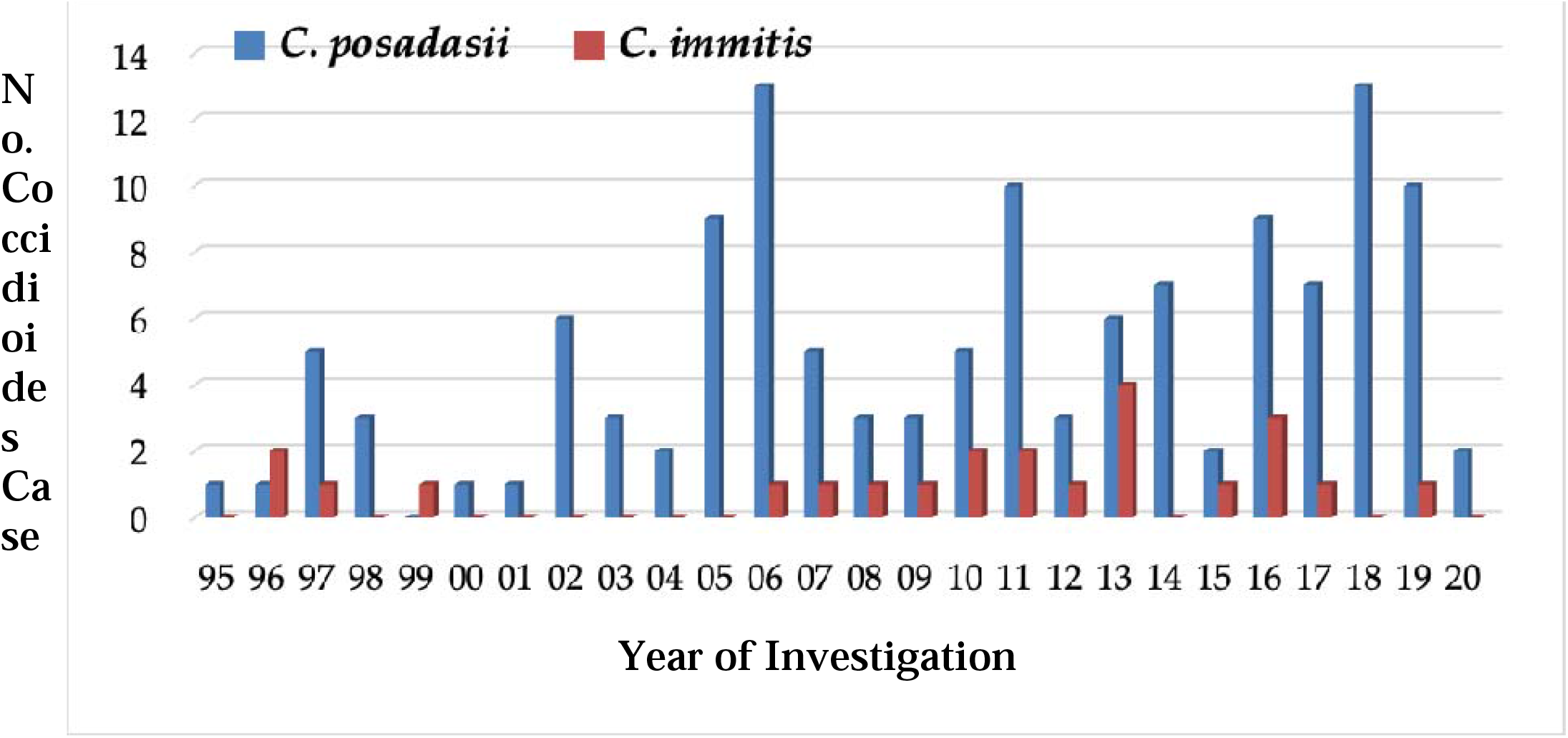
Human coccidioidomycosis cases from 1995 to 2020. Isolates received from various facilities from New York and neighboring states were analyzed retrospectively by newly developed duplex real-time PCR assay. Of 152 cases, 129 were confirmed as being caused by *C. posadasii*, and 23 were caused by *C. immitis*. The predominance of *C. posadasii* over *C. immitis* observed in the isolates analyzed.

## DISCUSSION

PCR-based methods have been used increasingly due to their accuracy, sensitivity, and speed of identification, and the use of DNA instead of highly infectious live cultures. Microsatellite analysis has been the primary molecular method used to distinguish the two species of *Coccidioides* (11, 12). However, this method is both time consuming and technically challenging for clinical and reference laboratories. The light-cycler PCR assay developed by Binnicker et al. (17) was highly sensitive and specific for *Coccidioides* species, but this assay could not distinguish *C. immitis* from *C. posadasii*. Umeyama et al. (15) described disparity PCR using species-specific primers that resulted in different size amplicons for a convenient distinction of *C. immitis* from *C. posadasii* by conventional PCR. In the present study, we modified the Umeyama et. al. (15) disparity PCR, which led to the reduction in amplicon size of 200-bp for *C. immitis* and 114-bp for *C. posadasii*. The conventional diagnostic PCR assay was excellent for the identification of *C. immitis* and *C. posadasii* from culture, but it was not good for the analysis of *Coccidioides* DNA from paraffin-embedded primary specimens. One reason could be that the amount of *Coccidioides* DNA present in these samples were below the threshold for detection by conventional PCR, or the template was degraded beyond the capacity of the conventional PCR to yield a positive result. PCR allelic-discrimination assay, referred to as CocciDiff, was developed using TaqMan chemistry (14). This assay could effectively identify *Coccidioides* cultures and could accurately distinguish *C. immitis* from *C. posadasii* based upon the presence of an individual highly informative canonical SNP (14). CocciDiff has yet to be validated with clinical specimens. Although, the CocciDiff assay appears to be quite promising, the assay specificity based on canonical SNP can be problematic if such mutations are not consistently present in a given strain of *Coccidioides*. In the present study, while developing a duplex real-time PCR assay, we used more stringent conditions where first set of primers and probe targeting *PRA2* gene identified *Coccidioides* to the genus level, and the second set of primers and probe targeting *C. immitis* contig, Ci41815 (CiC), identified *C. immitis* but not *C. posadasii* due to a deletion of 86-bp sequence in the corresponding Cp41810 contig. Since there are only two species within the *Coccidioides* genus, the positive results with both probes of the duplex assay allowed the identification of *C. immitis* while positive, negative results with probes *PRA2* and Ci41815, respectively, led to the identification of *C. posadasii*.

Of 129 *C. posadasii*, 117 isolates were from patients from New York, followed by eight isolates from patients from New Jersey, two isolates each from patients from Pennsylvania and Arizona, one isolate from a patient from Washington DC. All 23 *C. immitis* isolates were from patients from NY. Since *Coccidioides* is not found in the soil of New York and other states in the northeast region, the positive isolates suggested that the patients acquired *Coccidioides* infection while travelling to the area of endemicity. These findings are consistent with our earlier study where travel history to endemic areas for *Coccidioides* was linked to positive clinical cases from New York (18). In the present study, we confirmed five-fold more isolates as *C. posadasii* than as *C. immitis*; this observation supports a previous publication on possible larger population size and more diverse distribution of *C. posadasii* vis-a-vis *C. immitis* (19). Among a smaller sampling of isolates from California, we found four *C. posadasii* isolates. Also, positive RhMK cell samples came from a monkey housed in California were also identified as *C. posadasii*. These results raised the possibility that a small niche of *C. posadasii* exists in California or patients might have traveled to areas where *C. posadasii* is endemic. Further studies are needed to delineate the geographic distribution of *C. immitis* and *C. posadasii* and the areas of overlap if any.

Serology tests are the mainstay of coccidioidomycosis diagnosis (20, 21). Our study included a small sample size of serology-positive clinical specimens, but the findings do not permit firm correlation with real-time PCR test. The gold standard for the diagnosis of coccidioidomycosis is the culture of the organism from primary specimens. Culture is highly sensitive, and the DNA probe for confirmatory testing of culture isolates has yielded excellent specificity (22, 23). However, growth in culture may take several days to weeks resulting in a delay in diagnosing and initiating treatment in infected individuals. The highly infectious nature of arthrospores produced by *Coccidioides* species presents a safety risk to the laboratory personnel if a culture is not quickly identified and handled appropriately under biosafety level 3 containment. It is important to note that laboratory-acquired infections due to *Coccidioides* species have been reported in the literature (24). In summary, our modified diagnostic conventional PCR assay was excellent for the culture identification of *C. immitis* and *C. posadasii*, and the new duplex real time PCR assay had broader utility for identifying two *Coccidioides* species from culture and primary specimens. The *Coccidioides* spp. duplex real-time PCR will allow rapid differentiation of *C. immitis* and *C. posadasii* from clinical specimens and further augment coccidioidomycosis surveillance.

## Data Availability

The data will be available referred in the manuscript

## ACKNOWLEDGEMENTS

We acknowledge the Wadsworth Center (WC) Tissue Culture & Media, and the Applied Genomic Technologies Cores for media and sequencing services. Dr. Wallace, Milwaukee Zoo, thanked for providing Rhino tissues suspected of coccidioidomycosis. Dr. Kimberlee McClive-Reed provided editorial comments on the draft manuscript. We thank YanChun Zhu for assistance with the reference testing of *Coccidioides* spp. VC thanks Dr. Edward Desmond and other staff members at the California Department of Public Health for assistance with implementation of *Coccidioides* real-time PCR test. This work was supported partly by funds from the WC, the New York State Department of Health (NYSDOH), and the Centers for the Disease Control and Prevention (CDC) grant number NU50CK000516. The contents of this manuscript are solely the responsibility of the authors and do not necessarily represent the official views of the NYSDOH or the CDC.

## AUTHOR CONTRIBUTIONS

SC conceived the study, designed primers and probes for duplex and conventional PCR assays, supervised experiments, and wrote the manuscript. TV, AM, and KS standardized, validated single-plex, duplex real-time, and conventional PCR assays. KLC contributed to the clinical specimen selection, helped in data interpretation, and approved final draft. VC contributed to the study design, procured *Coccidioides* isolates and primary specimens from other agencies and edited the draft manuscript.

## FIGURE LEGEND

**Supplementary Table 1.** Specificity Panel.

| **Supplementary Table 1. Specificity Panel** |  |  |  |  |
| --- | --- | --- | --- | --- |
| **ISOLATE No.** | **ORGANISM** | **SOURCE** | **PRA2** | **CiC** |
| - | NTC | - | NEGATIVE | NEGATIVE |
| **Closely related mold species** |  |  |  |  |
| M808 | *Blastomyces dermatitidis* | MCC, NYSDOH | NEGATIVE | NEGATIVE |
| M809 | *Blastomyces dermatitidis* | MCC, NYSDOH | NEGATIVE | NEGATIVE |
| 18461-12 | *Blastomyces dermatitidis* | MCC, NYSDOH | NEGATIVE | NEGATIVE |
| 20694-12 | *Histoplasma capsulatum* | MCC, NYSDOH | NEGATIVE | NEGATIVE |
| 22276-12 | *Histoplasma capsulatum* | MCC, NYSDOH | NEGATIVE | NEGATIVE |
| M109 | *Sporothrix schenckii* | MCC, NYSDOH | NEGATIVE | NEGATIVE |
| K10 | *Sporothrix schenckii* | MCC, NYSDOH | NEGATIVE | NEGATIVE |
| 12998-12 | *Chrysosporium* sp. | MCC, NYSDOH | NEGATIVE | NEGATIVE |
| M1512 (K25) | *Chrysosporium* sp. | MCC, NYSDOH | NEGATIVE | NEGATIVE |
| M1542 (K33) | *Paracoccidioides brasiliensis* | PHL, U.K." | NEGATIVE | NEGATIVE |
| **Distantly related mold species** |  |  |  |  |
| 17452-12 | *Trichophyton rubrum* | MCC, NYSDOH | NEGATIVE | NEGATIVE |
| K2 | *Trichophyton mentagrophyte* | MCC, NYSDOH | NEGATIVE | NEGATIVE |
| 19616-12 | *Trichophyton interdigitale* | MCC, NYSDOH | NEGATIVE | NEGATIVE |
| K3 | *Microsporum gypseum* | MCC, NYSDOH | NEGATIVE | NEGATIVE |
| 1304101 | *Geomyces pannorum* | MCC, NYSDOH | NEGATIVE | NEGATIVE |
| K5 | *Geomyces destructans* | MCC, NYSDOH | NEGATIVE | NEGATIVE |
| 16341-12 | *Aspergillus fumigatus* | MCC, NYSDOH | NEGATIVE | NEGATIVE |
| 15817-12 | *Aspergillus versicolor* | MCC, NYSDOH | NEGATIVE | NEGATIVE |
| 15822-12 | *Aspergillus terreus* | MCC, NYSDOH | NEGATIVE | NEGATIVE |
| 13071-12 | *Aspergillus nidulans* | MCC, NYSDOH | NEGATIVE | NEGATIVE |
| 12745-12 | *Wangiella dermatitidis* | MCC, NYSDOH | NEGATIVE | NEGATIVE |
| 10461-12 | *Penicillium pururogenm* | MCC, NYSDOH | NEGATIVE | NEGATIVE |
| 13041-12 | *Penicillium funiculosum* | MCC, NYSDOH | NEGATIVE | NEGATIVE |
| 13446-12 | *Fusarium spp* | MCC, NYSDOH | NEGATIVE | NEGATIVE |
| M831 (K12) | *Epidermophyton floccosum* | MCC, NYSDOH | NEGATIVE | NEGATIVE |
| **Distantly related yeast species** |  |  |  |  |
| NIH444 | *Cryptococcus gattii* | ATCC90028 | NEGATIVE | NEGATIVE |
| NIH12 | *C. neoformans* var. *neoformans* | MCC, NYSDOH | NEGATIVE | NEGATIVE |
| H99 | *C. neoformans* var. *grubii* | MYCC, NYSDOH | NEGATIVE | NEGATIVE |
| K16 | *C. albidus* | MYCC, NYSDOH | NEGATIVE | NEGATIVE |
| M49 (K17) | *Candida albicans* | ATCC22019 | NEGATIVE | NEGATIVE |
| K19 | *Candida tropicalis* | ATCC750 | NEGATIVE | NEGATIVE |
| K18 | *Candida parapsilosis* | ATCC22019 | NEGATIVE | NEGATIVE |
| K19 | *Candida tropicalis* | ATCC750 | NEGATIVE | NEGATIVE |
| M146 | *Candida krusei* | ATCC6258 | NEGATIVE | NEGATIVE |
| M165 | *Candida glabrata* | MCC, NYSDOH | NEGATIVE | NEGATIVE |
| M157 | *Candida dubliniensis* | MCC, NYSDOH | NEGATIVE | NEGATIVE |
| M277 | *Geotrichum candidum* | MCC, NYSDOH | NEGATIVE | NEGATIVE |
| 771-07 | *Trichosporon asahii* | MCC, NYSDOH | NEGATIVE | NEGATIVE |
| M161 (K22) | *Rhodotorula mucilaginosa* | MCC, NYSDOH | NEGATIVE | NEGATIVE |
| M78 (K24) | *Saccharomyces cerevisiae* | MCC, NYSDOH | NEGATIVE | NEGATIVE |
| **Bacterial Species** |  |  |  |  |
| S-1 | *Mycoplasma pneumoniae* | BCC, NYSDOH | NEGATIVE | NEGATIVE |
| S-2 | *Pseudomonas aeruginosa* | BCC, NYSDOH | NEGATIVE | NEGATIVE |
| S-3 | *Nocardia farcinica* | BCC, NYSDOH | NEGATIVE | NEGATIVE |
| S-4 | *Streptococcus pneumoniae* | BCC, NYSDOH | NEGATIVE | NEGATIVE |
| C-735 (Co) | *C. posadasii* | MCC, NYSDOH | POSITIVE | POSITIVE |
| MYC249-09 (Co) | *C. immitis* | MCC, NYSDOH | POSITIVE | POSITIVE |

ATCC = American Type Culture Collection; MCC = Mycology Culture Collection, New York State Department of Health (NYSDOH); BCC = Bacterial Culture Collection (NYSDOH), PHL, U.K; Public Health Laboratory, United Kingdom

**Supplementary Table 2A.** Inter-Assay Reproducibility.

| Supplementary Table 2A- Inter-Assay Reproducibility |  |  |  |  |  |  |  |
| --- | --- | --- | --- | --- | --- | --- | --- |
| Organisms | Isolate<br>No. (ng/PCR<br>rxn) | <u>PRA2-Cy5</u> |  |  | <u>CiC-FAM</u> |  |  |
|  |  | Day 1<br>Ct1/Cy2 | Day 2<br>Ct1/Cy2 | Day 3<br>Ct1/Ct2 | Day 1<br>Cy1/Ct2 | Day 2<br>Ct1/Ct2 | Day 3<br>Ct1/Ct2 |
| *C. immitis* | 13-7701(10) | 22.61/22.78 | 22.67/22.80 | 22.05/21.98 | 22.04/21.96 | 21.60/21.91 | 22.59/22.20 |
|  | 13-2789 (10) | 23.81/23.65 | 23.31/23.53 | 23.94/23.85 | 22.04/21.96 | 22.99/23.17 | 22.54/22.78 |
|  | 664-98 (0.1) | 29.45/29.45 | 30.56/30.25 | 30.23/31.84 | 30.15/30.39 | 30.09/30.40 | 30.84/30.09 |
| *C. posadasii* | 14-1599 (100) | 18.84/18.99 | 18.70/18.68 | 19.37/19.89 | 0.0 | 0.0 | 0.0 |
|  | 13-2793 (10) | 24.03/23.82 | 23.61/23.67 | 22.53/22.43 | 0.0 | 0.0 | 0.0 |
|  | 13-4872 (1) | 27.81/27.70 | 27.00/26.96 | 26.90/29.29 | 0.0 | 0.0 | 0.0 |

**Supplementary Table 2B.** Intra-Assay Reproducibility.

| Supplementary Table 2B- Intra-Assay Reproducibility |  |  |  |  |  |  |  |
| --- | --- | --- | --- | --- | --- | --- | --- |
| Organisms | Isolate<br>No. (ng/PCR rxn) | <u>PRA2-Cy5</u> |  |  | <u>CiC-FAM</u> |  |  |
|  |  | Ct1 | Ct2 | Ct3 | Ct1 | Ct2 | Ct3 |
| *C. immitis* | 13-7701 (1) | 24.81 | 24.52 | 24.44 | 24.66 | 24.60 | 24.68 |
|  | 13-2789 (10) | 22.55 | 22.66 | 22.58 | 22.49 | 22.43 | 22.54 |
|  | 664-98 (0.1) | 29.97 | 29.77 | 29.79 | 29.49 | 30.65 | 30.84 |
| *C. posadasii* | 14-1599 (100) | 19.37 | 19.89 | 19.84 | 0.0 | 0.0 | 0.0 |
|  | 13-2793 (10) | 22.53 | 22.43 | 22.84 | 0.0 | 0.0 | 0.0 |
|  | 13-4872 (1) | 26.23 | 26.50 | 26.90 | 0.0 | 0.0 | 0.0 |

**Supplementary Table 4.**
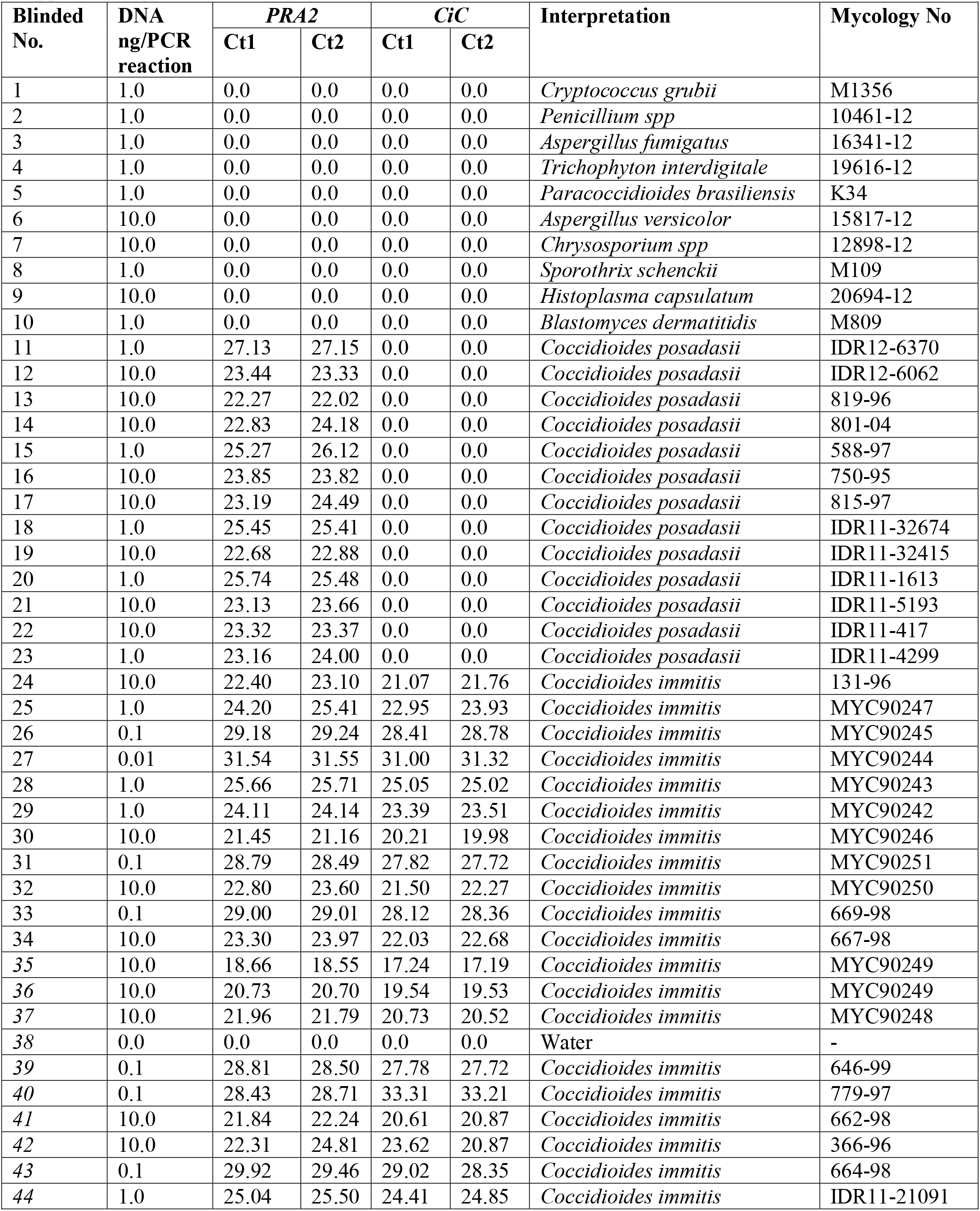
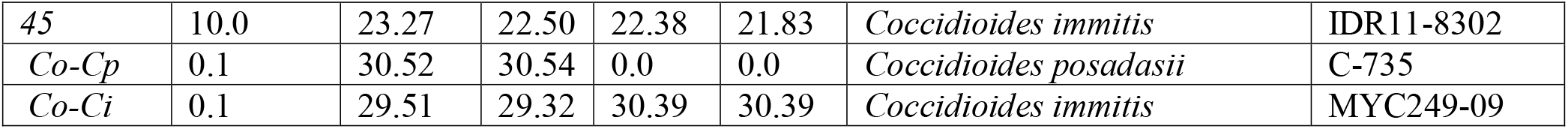
Blinded Panel.

## Notes

### Competing Interest Statement

The authors have declared no competing interest.

### Clinical Trial

Not applicable

### Author Declarations

1.Christina Egan, Ph.D. FROM: Robin Krause, IRB Administrator I New York State Department of Health Institutional Review Board The study was exempt from IRB approval as residual samples were used for the investigation for which Wadsworth Center has approval.

